# Triangulating the neural cornerstones of reading: Within-participant double dissociations induced by direct cortical stimulation

**DOI:** 10.1101/2025.04.13.25324235

**Authors:** Masako Daifu-Kobayashi, Akihiro Shimotake, Makiko Ota, Mitsuhiro Sakamoto, Katsuya Kobayashi, Takayuki Kikuchi, Kazumichi Yoshida, Takeharu Kunieda, Susumu Miyamoto, Ryosuke Takahashi, Akio Ikeda, Matthew A. Lambon Ralph, Riki Matsumoto

## Abstract

How does the human brain recognize a written word and read it aloud? The “triangle” neurocognitive model proposes that reading is founded on three cognitive systems: visual-orthography, phonology and semantics. We tested this theory via a unique opportunity to generate transient double dissociations within the same patient via direct cortical stimulation to contrastive regions. Five patients with intractable temporal lobe epilepsy who underwent presurgical evaluation with subdural electrode were included. Reading of kanji words, with three levels of spelling-to-sound consistency level, and kana non-words with 2 to 5 mora, was examined. Low-intensity electrical stimulation was applied to the anterior basal temporal language area (BTLA) (<6 cm from the temporal pole, all patients), posterior BTLA (>6 cm, in four patients) and supramarginal gyrus (SMG, one patient). Electrical or sham stimulation was delivered time-locked to the presentation of each word, and the reaction time (RT) was evaluated. One double dissociation was generated by contrastive SMG (phonological alexia; poor non-word reading) vs. anterior BTLA stimulation (surface dyslexia; poor kanji reading). A different double dissociation was elicited following anterior vs. posterior BTLA stimulation with the former showing effects of kanji consistency level as expected in surface dyslexia, and the latter with length effects as expected in a pure alexia. In conclusion, going beyond classic lesion studies, we were uniquely able to demonstrate the double dissociations within the same patient on a transient reversible basis. Consistent with the triangle model of reading, three different reading disorders were elicited following low-intensity stimulation to each primary cognitive system.

## Introduction

The ability to read is fundamental to education, professional activities and everyday pursuits, and has increased its importance since the rise of the internet, email and social media for our sources of knowledge exchange. Accordingly, a central neuroscience question asks: what are the neural bases and associated cognitive-language processes that allow the human brain to recognize a written word and to read it aloud? Amongst the different types of language activity that we are able to engage in, reading is unique in that it requires the interaction of the brain’s fundamental language and visual systems. Ever since the rise of classical 19th century cognitive neurology, this question has been explored through neuropsychological studies of patients with chronic reading impairments (1–3) and in contemporary times through functional neuroimaging and transcranial magnetic stimulation (TMS) in healthy participants (4–7). In this study, we report insights afforded by the rare opportunity to investigate the transient effects of direct cortical stimulation in human neurosurgical patients. Unlike other neuroscience methods, this approach offers multiple research advantages including the ability to explore performance quantitatively before, during and after stimulation; and, when stimulating different key brain areas, the emergence of transient dissociation and double dissociations within the *<u>same</u>* patient (i.e., each patient is their own control, which is impossible in traditional lesion-based neuropsychology).

The study was motivated by the neurocognitive framework for reading proposed under the “primary systems hypothesis” (8) and encapsulated formally though the “triangle” computational models of reading (9–11). This framework proposes that three “primary” cognitive systems (visual-extraction of orthographic information, phonology and semantics) are the foundation of reading. Through their interconnection, two major processing pathways are formed: (a) the ‘direct’ dorsal pathway — direct translation of orthography to phonology; and (b) the ‘indirect’ semantic, ventral pathway — a semantically-mediated pathway (3, 10). The direct pathway contributes to the pronunciation of all types of word and non-word, whereas the indirect semantic pathway is involved only when reading words which are known to a reader. This indirect pathway is especially important for reading words with atypical pronunciations given that the direct pathway tends to generate the orthographic-to-phonologically typical pronunciation (3).

To date, the contribution of each primary system to reading has been explored separately, which we addressed in this investigation through parallel investigation within the same participants (before and during direct stimulation of contrastive cortical sites). A core source of evidence for the contribution of the anterior temporal lobe (ATL) semantic system to reading, comes from patients with semantic dementia (SD). In this neurodegenerative disorder, patients show a selective yet progressive degradation of multimodal semantic memory consequent on atrophy centered on the ventrolateral ATL, bilaterally. (12–14). In line with the computational models of reading, this progressive semantic decline is almost always accompanied by surface dyslexia in which patients show inaccurate reading of low frequency words with atypical spelling-to-sound pronunciations and generate regularization errors (3). Convergent evidence for role of the ATL semantic hub in reading aloud has been derived in healthy participants from targeted distortion-minimising functional magnetic resonance imaging (fMRI) (15) and repetitive transcranial magnetic stimulation (rTMS) (7, 16). These studies indicate that the ventral ATL (vATL) is a key subregion, an area that aligns with the anterior part of the BTLA as highlighted by seminal functional cortical stimulation mapping (17, 18). Indeed, not only have fMRI and rTMS studies have also confirmed this as an important semantic region (19) but also contemporary cortical-grid stimulation and decoding of electrocorticogram (ECoG) data have demonstrated semantic coding in the surface of the ventral ATL (20, 21, 22). Based on these findings, the indirect semantic reading pathway in the triangle model is assumed to go through the anterior part of BTLA.

The posterior BTLA extends to the ventral occipitotemporal region which includes the visual word form area (VWFA) (23–25). Although the exact mechanisms of this region are debated, it is clear from lesion studies (26, 27), functional neuroimaging and rTMS in health participants (28, 29), and a cortical stimulation mapping study (30) that the ventral occipito-temporal (vOT) region is important for extracting orthographic and word information from visual input.

Finally, phonological processing appears to be supported by a network of brain regions including inferior prefrontal cortex, posterior superior temporal gyrus (STG) as well as the inferior SMG (31). Many neural models of reading assume that the SMG provides a nexus for the connection from visual areas into the phonological network (32, 33). Consistent with these proposals, the SMG has been shown via fMRI and rTMS to be engaged when healthy participates make phonological decisions (5, 6, 34–38) and, when damaged, it can give rise to phonological impairments (conduction aphasia) (39–41) and phonological alexia (42).

The present cortical stimulation study explored the effect of stimulation to three different regions on reading Japanese. This also allowed us to explore the unique features of Japanese orthographic systems. Briefly, the Japanese writing system has two distinct orthographies, kanji and kana. Kanji is a semi-opaque orthography and its characters are strongly associated with semantics. Kana (written in either hiragana or katakana) has the characteristic that it has only one pronunciation for each kana character (each corresponds to a single Japanese syllable or mora). Accordingly, Japanese SD patients show poor reading of words written in kanji more than in hiragana (43, 44). In contrast, Japanese patients with phonological alexia show the typical symptom of a lexicality effect with better reading of words (written in either script) than non-words (45).

The current study explored the contrastive effects of transient cortical stimulation to each of these three regions; with each patient providing data on reading before and during stimulation in two of these areas. The simulation of multiple areas within the same patient meant that, unlike traditional lesion-based neuropsychology, we were able to generate transient double dissociations within each patient and also ensure that their baseline performance was normal. We used the same stimulation approach as that used previously to confirm the semantic characteristics of the vATL (46). Our study was split into three investigations: (1) contrasting vATL (semantic ventral pathway) vs. SMG (dorsal phonological direct pathway) stimulation which generated transient surface vs. phonological alexia (respectively) — a pattern that as far as we are aware has never been shown within the same individual before; (2) vATL vs. posterior-mid BTLA which generated contrastive reading patterns consistent with the contributions of semantics (surface dyslexia) vs. visual/orthography processing (pure alexia) to reading; (3) finally, recent explorations of reading in English SD patients (3), and TMS and fMRI studies in healthy participants (15, 16) suggest that the relative contribution of each reading pathway varies across individuals. Accordingly, we undertook an exploration of the effects of individual differences in semantic reliance (SR) on reading typical and atypical kanji words.

## Results

### 1. Double dissociation of the ventral and dorsal reading pathway (Patient 5)

We evaluated the effect of direct cortical stimulation on reading kanji words with different consistency levels and kana non-words with different mora lengths (see Fig. 5 for detailed methods). In Patient 5, we stimulated an electrode pair located in the anterior BTLA vs. in SMG (Figure 1, A). During anterior BTLA stimulation, there was a clear stimulation effect for kanji word reading but no effect on kana non-word reading [the stimulation effect was greater on kanji reading than kana non-words reading; F = 17.510, p < 0.001] (Figure 1, B, lower panel). Moreover, the anterior BTLA stimulation generated a pattern consistent with surface dyslexia in Japanese orthographies (replicated in the larger patient sample reported below): specifically, a two-way ANOVA with the factors of kanji type (C: consistent, IT: inconsistent typical, IA: inconsistent atypical) and stimulation (sham, stimulation) revealed a significant interaction between kanji type and stimulation (F = 3.347, p = 0.04; Figure 1, C, lower panel). As expected, the vATL stimulation had a much greater effect on IA word reading than in C words (F = 6.457, p = 0.014). In contrast and again as expected for surface dyslexia, the same vATL stimulation had no effect on non-word reading. Specifically, a two-way ANOVA with the factors mora length (2–3 mora, 4–5 mora) and stimulation (sham, stimulation) did not show any main effects (mora: F = 1.475, p = 0.23, stimulation: F = 0.244, p = 0.624) or interaction between mora length and stimulation (F = 0.328, p = 0.570) (Figure 1, D, lower panel).

**Figure 1.**
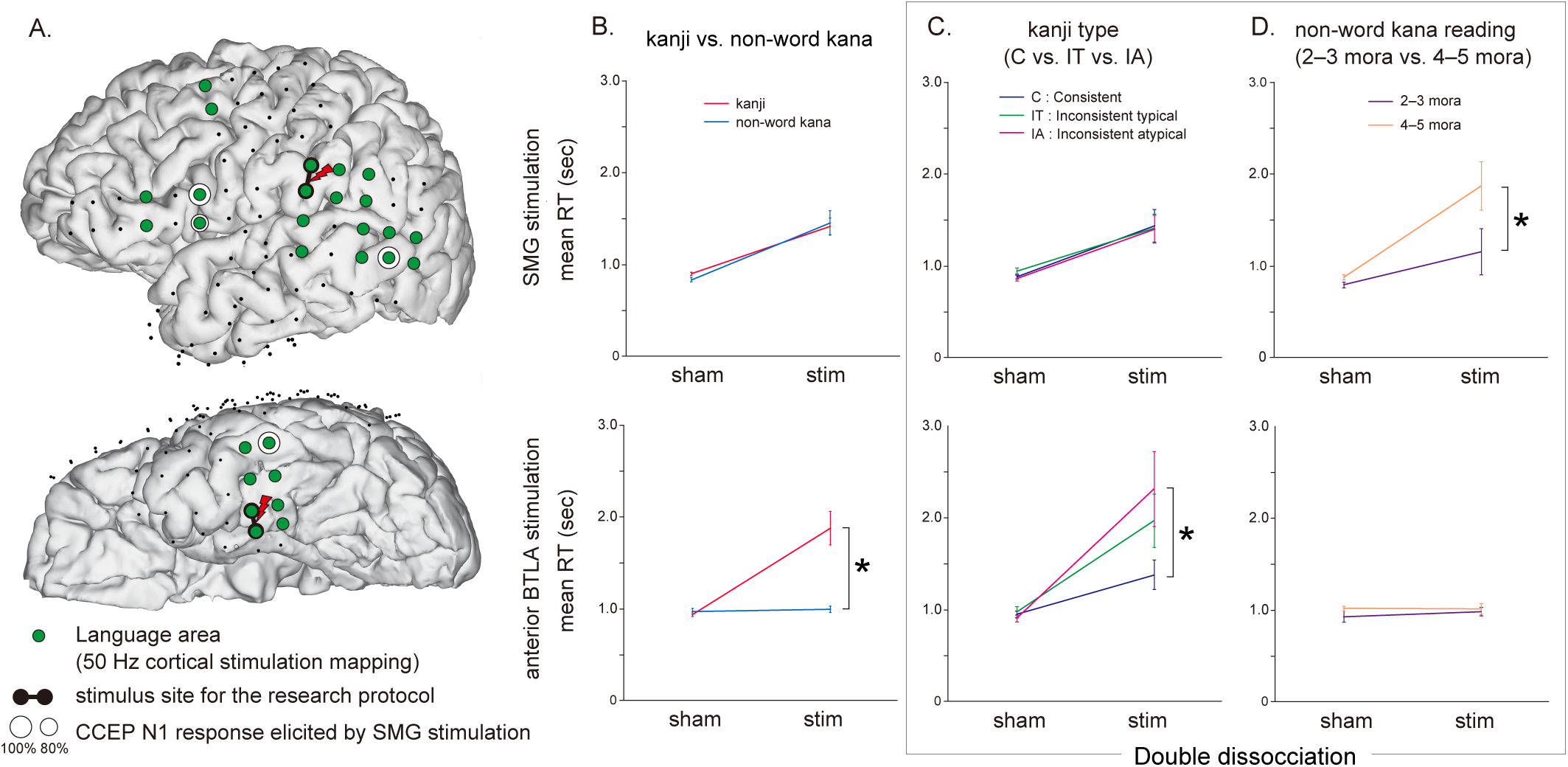
Double dissociation of kanji and kana reading between supramarginal gyrus (SMG) and anterior basal temporal language area (BTLA) in Patient 5. A. We stimulated the anterior BTLA and the SMG (red lightning shape). The green circles are the electrodes which showed language function by 50 Hz electrical cortical stimulation mapping for clinical purpose. Single-pulse electrical stimulation at SMG elicited the early negative response (N1) of cortico-cortical evoked potential (CCEP) in the remote language areas including Broca’s area, indicating that the site of stimulation at SMG is a part of the dorsal language pathway. B. The reaction time (RT) of kanji and non-word kana reading during the SMG stimulation (sham and stim) and the anterior BTLA stimulation (sham and stim). C. The RT of kanji reading with each kanji types (C: Consistent, IT: Inconsistent typical, IA: Inconsistent atypical) during the SMG and the anterior BTLA stimulation (sham and stim). The RT of consistent (C) words reading is shown as a blue line, inconsistent typical (IT) words as a green line, and inconsistent atypical (IA) words as a pink line. D. The RT of non-word kana reading during the SMG and the anterior BTLA stimulation (sham and stim). The RT of 2–3 mora non-word kana is shown as a purple line, and 4–5 mora non-word kana as an orange line. Asterisks indicate significant interaction between task and stimulation.

The SMG stimulation in the same patient generated an opposite pattern, consistent with that expected for phonological alexia. During the SMG stimulation, the effect of the stimulation was almost the same between kanji reading and kana non-word reading: the stimulation affected both kanji and kana non-word reading (F = 55.624, p< 0.001) and no interaction was observed between stimulation and words types (F = 0.423, p = 0.516) (Figure 1, B, upper panel). The stimulation to the SMG slowed both kanji and kana non-words reading. Consistent with phonological alexia, SMG stimulation did not differentially affect the different types of kanji words but was sensitive to the length of the non-words (a hallmark of phonological impairments in phonological alexia) (47). Formally, the two-way ANOVA with the factors of SMG stimulation (sham, stim) and kanji type (C: consistent, IT: inconsistent typical, IA: inconsistent atypical) showed a main effect of stimulation (F = 34.654, p < 0.001) but no interaction between kanji type and stimulation (F = 0.093, p = 0.911; Figure 2, C, upper panel). The two-way ANOVA with the factors of SMG stimulation (sham, stim) and non-word mora length (2–3 mora, 4–5 mora) revealed a significant interaction between mora length and stimulation (F = 8.184, p = 0.006; Figure 1, D, upper panel), with a greater effect of SMG stimulation on 4–5 mora than 2–3 mora non-word reading.

**Figure 2.**
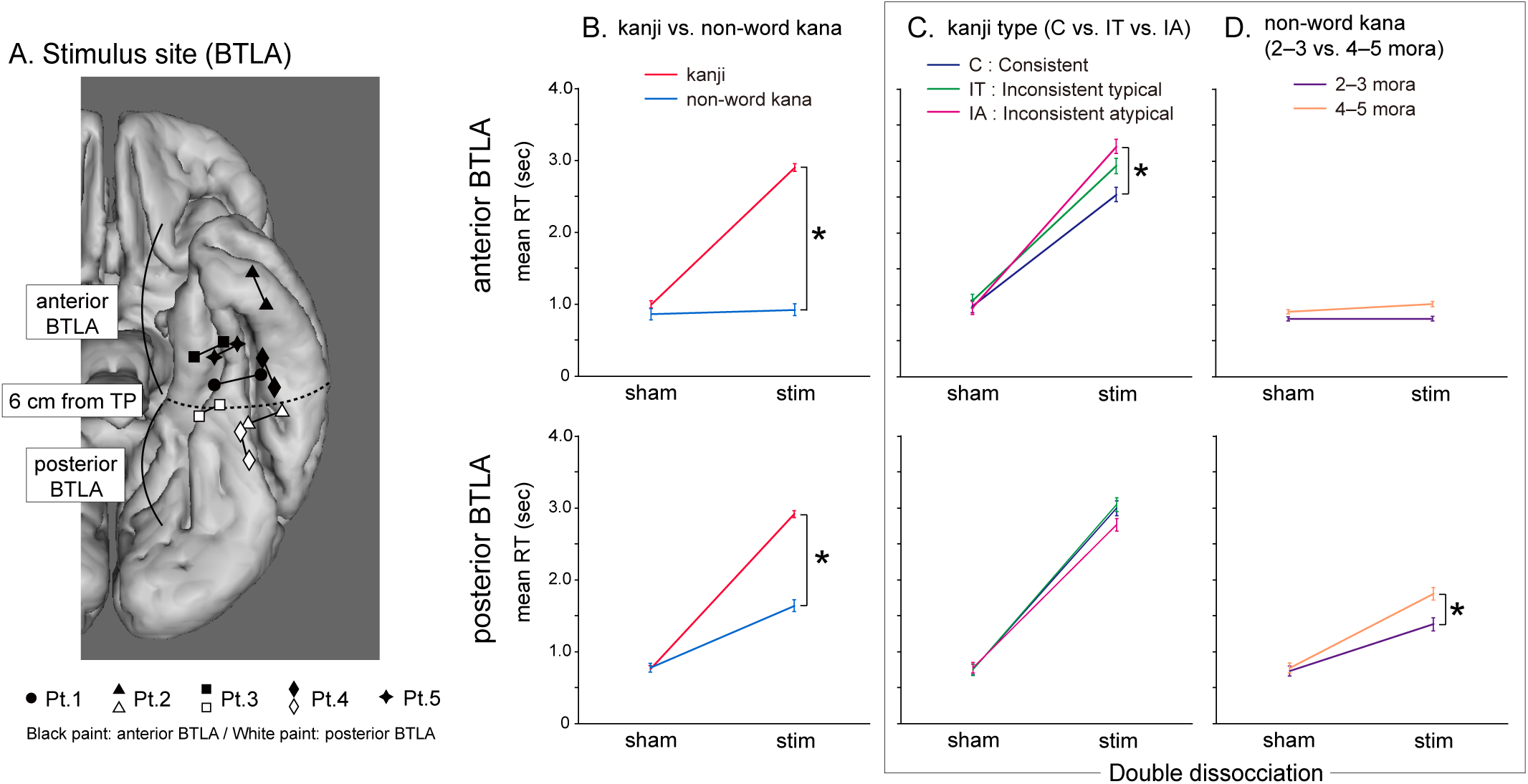
Double dissociation of kanji and kana reading between the anterior and posterior basal temporal language area (BTLA). A. Stimulus sites in the BTLA. We defined the anterior BTLA as falling within 6 cm of the tip of the temporal pole and the remaining posterior part was defined as the posterior BTLA. The electrodes with black paint were classified into the anterior BTLA and the electrodes with white paint were classified into the posterior BTLA. Patient 2, 3 and 4 were stimulated with both anterior and posterior BTLA. Patient 1 was stimulated only with the anterior BTLA. Patient 5 was stimulated with the anterior BTLA and supramarginal gyrus (see Figure. 1). B. The reaction time (RT) of whole kanji and non-word kana reading during the anterior and posterior BTLA stimulation (sham and stim). C. The RT of kanji reading with each kanji types (C: Consistent, IT: Inconsistent typical, IA: Inconsistent atypical) during the anterior and posterior BTLA stimulation (sham and stim). D. The RT of non-word kana reading during the anterior and posterior BTLA stimulation (sham and stim). The colors of the lines are the same as Figure 1. Asterisks indicate significant interaction between task and stimulation.

This strong double dissociation was reflected not only in the formal reading data (Figure 1, C vs. D) but also strikingly in Patient 5’s introspective comments. After anterior BTLA stimulation, Patient 5 complained that he could see and recognize the characters, but could not get the meaning of them. In addition, he knew the most typical ‘reading’ (orthographical to phonological conversion) of each kanji character but could not put the characters together and thus generate the correct (atypical) pronunciation for the whole word. In contrast after SMG stimulation, he complained that it was difficult to pronounce and read the words aloud.

### 2. Ventral reading pathway: Anterior vs. Posterior BTLA

We defined the anterior part of BTLA as falling within 6 cm of the tip of the temporal pole (in MNI space) and the remaining posterior part was defined as the posterior BTLA based on a previous cortical stimulation study (30) (Figure 2, A). As shown for Patient 5 individually, Patients 1–5 as a group showed the expected pattern for surface dyslexia after stimulation to the anterior BTLA (Figure 2, B, upper panel) with a strong effect of stimulation on kanji reading but absolutely no effect on kana non-word reading; formally, the by-items ANOVA showed a significant interaction between stimulation (sham, stim) and task (kanji vs non-word kana reading: F = 98.259; p < 0.001).

The pattern after stimulation to the posterior BTLA (Patients 2–4) (Figure 2, B, lower panel) was quite different and consistent with a more generalized effect as one would find in pure alexia, with an effect of stimulation on both kanji reading task (T =- 8.413, p < 0.01) and kana non-words reading task (T =-3.524, p < 0.01), with the ANOVA showing that the posterior BTLA stimulation had a greater effect on kanji than kana reading (a significant interaction between stimulation (sham, stim) and task (kanji vs non-word kana reading) (F = 99.553, p < 0.001).

We then looked at each reading task separately in order to explore the effect of stimulation on factors pertinent to each task. In line with previous studies of surface dyslexia in Japanese patients (44), we explored kanji words with different levels of orthographic-to-phonological consistency. Aligning with the pattern observed in surface dyslexia, after anterior BTLA stimulation, Patients 1–5 as a group showed an interaction between stimulation and kanji consistency level (see Figure 2, C, upper panel; F=6.032, p=0.003), with a greater effect of stimulation on IA words than C words (F=14.301, p<0.01). In contrast, after posterior BTLA stimulation (Figure 2, C, lower panel), there was an overall effect of stimulation (F = 889.585, p < 0.001) but no interaction with kanji type (F=2.032, p=0.133).

Finally, we explored the impact of anterior and posterior BTLA on reading of non-words with different lengths. Anterior BLTA stimulation, as found in surface dyslexia, had no effect on reading times (F = 1.957, p = 0.164) nor was there an interaction between mora length and stimulation (see Figure 2, D, upper panel; F = 2.382, p=0.125). In contrast and aligning with the length effects observed in pure alexia patients, not only was there a main effect of posterior BTLA stimulation (F = 92.819, p < 0.001) but there was also a significant interaction between mora length and stimulation (Figure 2, D, lower panel; F=5.237, p=0.024) reflecting the greater effect of stimulation on longer than shorter kana non-word reading.

Again, the formal empirical double dissociation observed after stimulation to the anterior vs. posterior BTLA (Figure 2, C vs. D) was mirrored in the patients’ introspections. For example, Patient 2 said that he had not been sure how to read the kanji characters and the meaning of them during the anterior BTLA stimulation. Whereas when stimulated in the posterior BTLA area, Patient 2 complained about the visual form, and read the kana words letter by letter partly. He said “I cannot see the whole word clearly”, “the letters look strange”, and “the letters are unclear”.

### 3. Preliminary exploration of individual differences in reading aloud after anterior BTLA stimulation – variation of semantic reliance (SR)

We conducted a preliminary investigation of the variations in effect of anterior BTLA stimulation on kanji word reading across the patients (Figure 3). A two-way ANOVA with kanji type (C: consistent, IT: inconsistent typical, IA: inconsistent atypical) and stimulation (sham, stimulation) revealed a significant interaction between kanji type and stimulation in Patient 1 (F = 4.092, p = 0.021), 4 (F = 3.460, p = 0.035) and 5 (F = 3.347, p = 0.040). The effect of stimulation on RT was greater for IA word reading than C word reading in Patient 1 and 5. For patient 4, a greater effect of stimulation was observed for IT word reading than C word reading. On the other hand, there was no interaction between Kanji type and stimulation in Patient 2 (F = 0.297, p = 0.744) and 3 (F = 1.226, p = 0.299).

**Figure 3.**
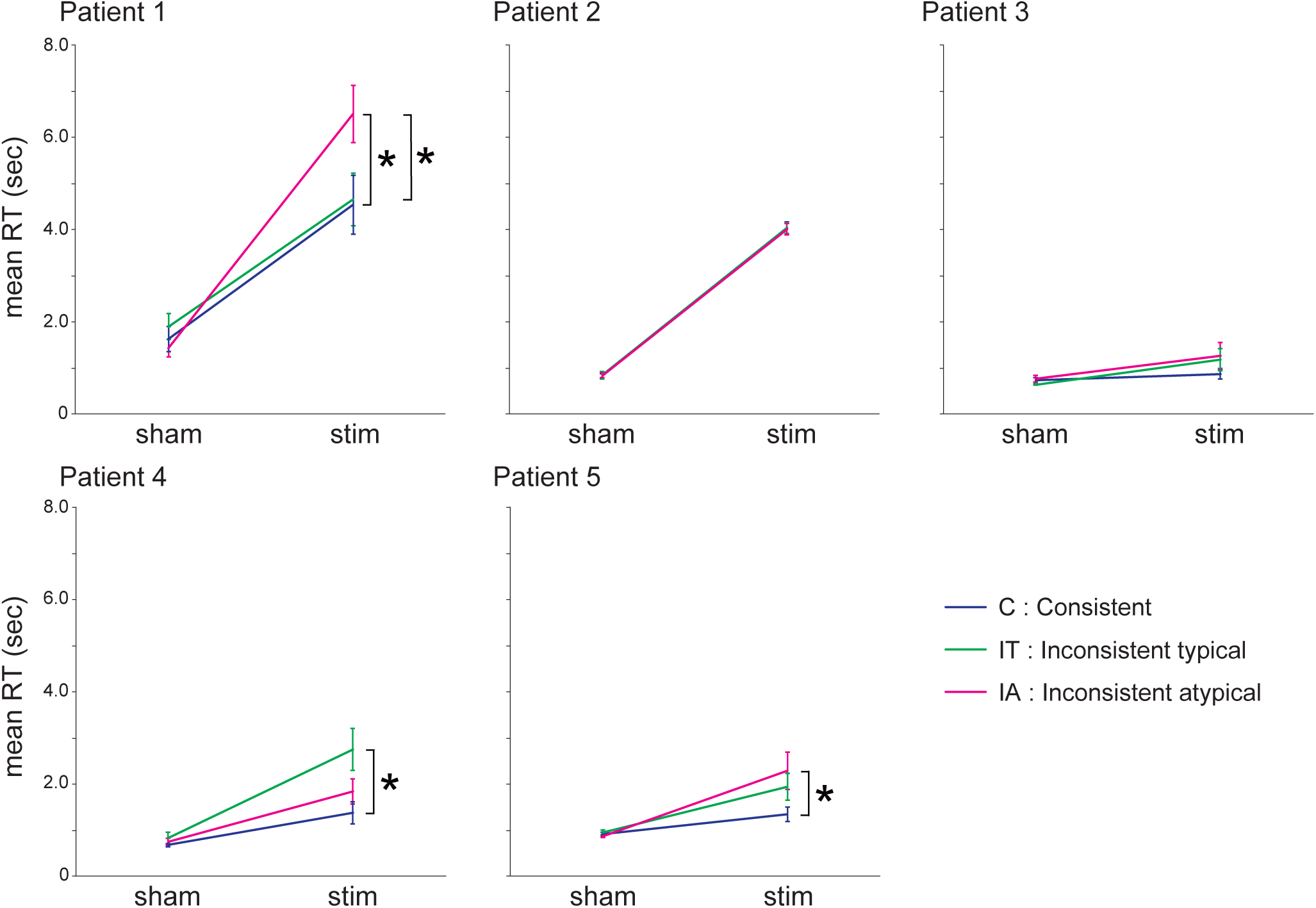
The individual reaction time (RT) of kanji reading with each kanji types (C: Consistent, IT: Inconsistent typical, IA: Inconsistent atypical) during the anterior basal temporal language area (BTLA) stimulation (sham and stim). The colors of the lines are the same as Figure 1 and 2. Asterisks indicate significant interaction between kanji type and stimulation. Patient 1, 4 and 5 showed significant interaction between kanji type and stimulation, while Patient 2 and 3 did not.

To evaluate if these individual differences might reflect variations in the SR of each patient (15, 16), we examined the level of correlation between kanji reading times from the presurgical baseline assessment and imageability, whilst partialing-out the effect of word frequency (Table SI-1). Patient 1 showed a significant partial correlation between reading speed and imageability in IA word reading (r = -0.54, p = 0.0058), while no correlation in C and IT word (C: r = -0.20, p = 0.25, IT: r = -0.28, p = 0.89). Patient 4 showed a trend-level partial correlation in IT word reading (r = -0.28, p = 0.091), while no correlation in C and IA word (C: r = -0.19, p = 0.25, IA: r = -0.17, p = 0.30). Patient 5 showed significant partial correlation in C word reading (r = -0.43, p = 0.0058) and a trend-level partial correlation in IT word reading (r = -0.31, p = 0.069), while no correlation in IA word (r = -0.18, p = 0.31). Patient 2 and 3 showed significant partial correlation or tendency of partial correlation in all items (Patient 2; C: r = -0.39, p = 0.015, IT: r = -0.38, p = 0.018 and IA: r = -0.58, p = 0.00011, Patient 3; C: r = -0.27, p = 0.094, IT: r = -0.43, p = 0.0078 and IA: r = -0.29, p = 0.091). Comparing these presurgical results with the effects of anterior BTLA stimulation during kanji word reading task, there was a correspondence between them except for Patient 5 (Figure 3 vs. Table SI-1). With the anterior BTLA stimulation during Kanji reading, Patient 1 showed consistency effect in IA, but not in C and IT. Patient 4 showed consistency effect in IT, but not in C and IA. Patient 2 and 3 showed no difference between all items.

## Discussion

In this study, we utilized the rare opportunity to investigate the transient effects of direct cortical stimulation in human neurosurgical patients. This approach offers the ability to explore performance quantitatively before, during and after stimulation; and, when stimulating different key brain areas, the emergence of transient double dissociations within the same patient. We used this powerful methodology to explore the neural bases of the triangle model (9–11) of reading (see Figure 4 through an exploration of Japanese and its two different scripts, kanji and kana. We were temporally able to create three different types of dyslexia (phonological, visual or surface) depending on where we stimulated. Across a case-series of patients we revealed two within-patient double dissociations for the first time: (a) stimulation to the anterior BTLA (associated with semantics) vs. SMG (associated with phonological processing) generated transient reading disorders consistent with surface vs. phonological alexia; and (b) a dissociation between the semantic and visual-orthographic aspects of reading by stimulation to the anterior vs posterior BTLA.

**Figure 4.**
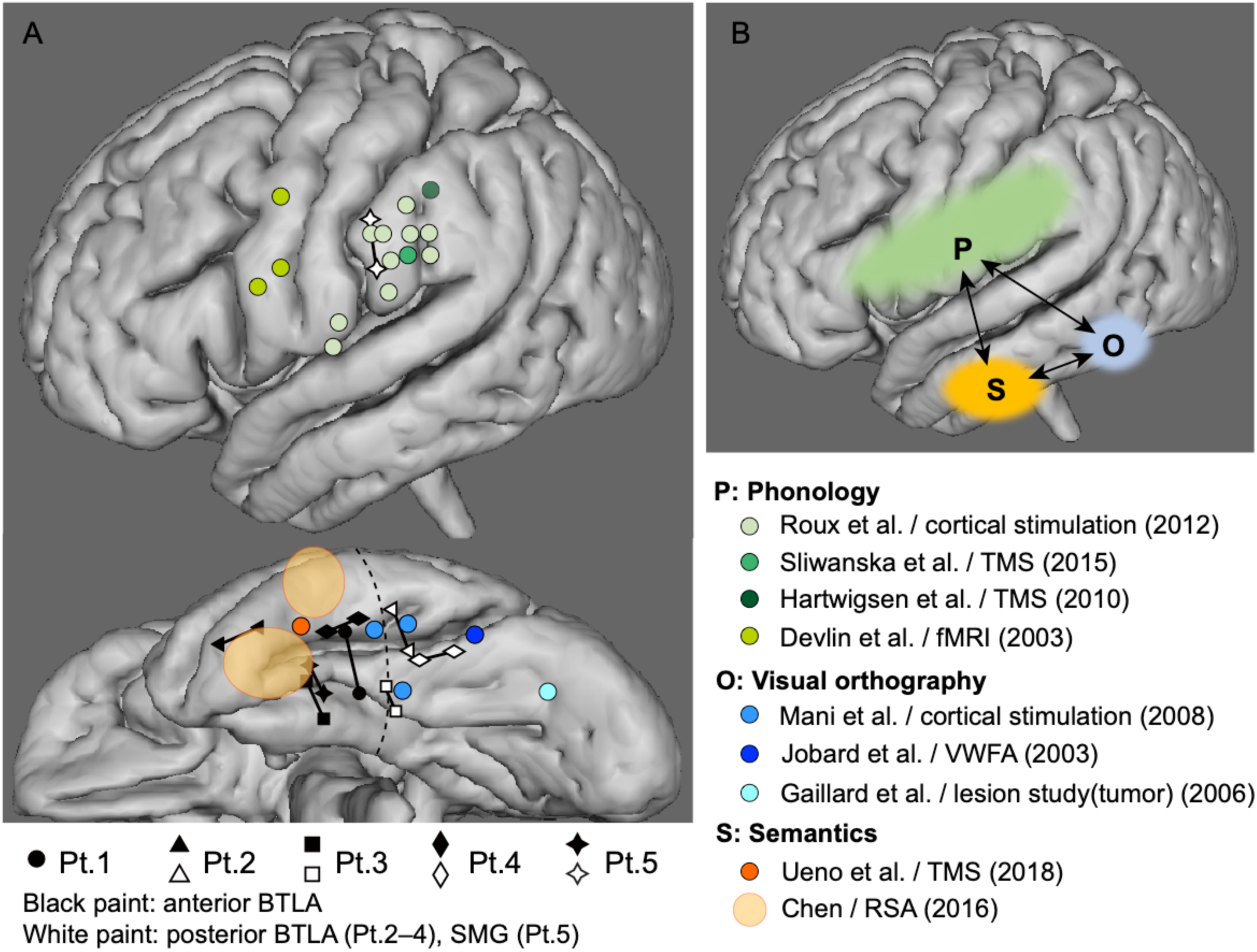
A. Comparison between reported studies and this study. The electrodes with black paint were classified into the anterior basal temporal language area (BTLA) and the electrodes with white paint were classified into the posterior BTLA in this study. The colored circles show the regions from previous stimulation studies and the temporal representational similarity analysis study. B. Schema of the triangle model on neuroanatomy, investigated in this study. S: semantic, P: phonology, O: orthography.

### Double dissociation of the anterior BTLA and SMG

During anterior BTLA stimulation, Patient 5 exhibited slowed reading for kanji; a pattern consistent with surface dyslexia as observed in patients with SD (consequent on atrophy centred on the anterior temporal lobe) (3, 44). We discuss these anterior BTLA results, alongside those found in the other BTLA-stimulated patients, in more detail below. In contrast after SMG stimulation, Patient 5 showed a pattern consistent with phonological alexia with greater slowing for non-word than word reading (mirroring the pattern observed in patients with phonological alexia after stroke) (42). The SMG electrodes stimulated in this study were consistent with the area stimulated by Sliwinska et al. (2012) (5) in their healthy-participant TMS study and highlighted the region’s importance to phonological processing. The area is also close to the region where impairment was observed during pseudoword reading in a cortical electrical stimulation study (38). The inferior parietal lobule, especially SMG is considered to be one of the important regions for phonological aspects of language based on functional neuroimaging (34, 35) and rTMS studies (5, 6, 36) (Figure 4). With regards to reading, Roux et al. (2012) (38) proposed that SMG performs the procedures necessary for converting spelling to sound during visual word recognition. Indeed, a wealth of neuropsychological studies have shown that this aspect of reading is heavily reliant on general phonological abilities (47, 48) for which SMG is one critical region (42). It is noteworthy that Patient 5 showed a word length effect during SMG stimulation, which is consistent with the phonological short term memory processes that have been associated with this region (49).

### Double dissociation between the anterior and the posterior BTLA

After stimulation to the anterior BTLA, kanji word but not kana reading times were slowed, and the patients exhibited an effect of kanji orthography-to-sound consistency, which is strikingly similar to the study of Japanese SD patients that employed the same kanji reading task (44). In contrast, stimulation to the posterior BTLA prolonged both kanji and kana reading times. Interestingly and unlike the anterior BTLA stimulation, a word length effect was observed in the kana reading task, while a consistency effect was not observed in kanji word reading. Indeed in addition to his introspective report of not seeing characters clearly, Patient 2 reported reading kana non-words character-by-character during posterior BTLA stimulation. A recent TMS study with Japanese healthy subjects used the same kanji task, and showed that TMS to an inevitably more lateral, vATL caused surface dyslexia (7). Another recent study, combining ECoG, fMRI and cortical stimulation mapping in epilepsy patients, showed that multimodal (auditory and visual) naming is reliant on the middle fusiform gyrus, which generally corresponds to the posterior portion of the anterior BTLA in our study (50).

The role of the left occipitoetemporal cortex in reading has been studied using a variety of methods including lesion studies, positron emission tomography (PET), fMRI, event-related potentials (ERPs) in intracranial recording, and cortical stimulation mapping (25–27, 30, 51). A single case study with a lesion in the left occipitotemporal area showed that a minute surgical resection caused letter-by-letter reading and selective disappearance of word-related fMRI activations. Based on this evidence, this region is considered to be involved in identifying words and letters, prior to activation of phonology or semantics. With regards to the role of this region in reading Japanese orthographies, a Japanese stroke study (52) reported that a hemorrhage lesion in the left posterior inferior temporal area caused impairment of kanji reading, while the caudal part of this region is involved in kana reading, and the rostral part is involved in naming. Previous functional neuroimaging studies (PET and fMRI) also showed that the left posterior inferior temporal area was activated during kanji reading (53, 54). Also in a study of a Japanese reading epilepsy patient, epileptic discharges were evoked in this area during katakana reading, which demands precise visual discrimination for word recognition (55).

In this study, we showed clear evidence for the first time that both kanji and kana reading are impaired during posterior BTLA stimulation in Japanese patients. Furthermore we revealed the functional division of labor between anterior and posterior BTLA in reading words, reflecting the double dissociation between semantics (anterior BTLA) and visual-orthographic processing (posterior BTLA) by means of direct electrical cortical stimulation.

### Semantic reliance (SR)

In this study, we also conducted a preliminary exploration of SR in each patient. Previous fMRI and TMS investigations of English reading have shown that SR and thus the engagement of the ATL semantic system, varies across healthy participants (15, 16) and leads to different levels of surface dyslexia in patients with SD (3). In line with these explorations, we also found that the SR of each patient varied such that a clearer pattern of kanji surface dyslexia emerged after anterior BTLA stimulation in the patients with the highest SR (gauged by the size of the imageability effect) in their baseline reading. Clinically, this is an important issue for considering the potential impact of ATL resection on semantic and language functions, and further studies, for example an evaluation of pre-surgical SR and influence on postoperative kanji reading ability, are necessary.

## Materials and Methods

### Participants

We recruited 5 patients with medically intractable temporal lobe epilepsy (TLE) who underwent chronic invasive recording with subdural electrodes which covered the language-dominant ventral part of the anterior temporal lobe before epilepsy surgery for identification of the seizure onset zone and eloquent cortices in Kyoto University Hospital from April 2014 to September 2017. The subdural electrodes were constructed of platinum with an interelectrode distance of 1 cm and a recording diameter of 2.3 mm (ADTECH, WI, USA). The patients’ demographics are summarized in Table SI-2. In all patients, the electrodes showing the earliest ictal ECoG changes, namely the seizure onset zone, were located on the parahippocampal gyrus, and on magnetic resonance imaging (MRI) all patients had hippocampal sclerosis in the language-dominant hemisphere. According to the Wada test, four out of five patients (Patient 1, 3–5) who had suffered from left TLE had language dominancy in the left hemisphere, and another one patient (Patient 2) with right TLE had language dominancy in the right hemisphere. All the five patients had normal language function as assessed by the Japanese version of the Western Aphasia Battery (WAB). Despite low verbal Intelligence Quotient (IQ) in Wechsler Adult Intelligence Scale III (WAIS-III) in Patient 1, we included her in this study because of her normal performance on the WAB.

This study was approved by the ethics committee of the Kyoto University Graduate School of Medicine (No. C533). Written informed consent was obtained from all the patients before the examination.

### Tasks (Kanji and non-word Kana)

The items used in this study were 120 kanji words consisting of two Chinese characters from Fushimi et al. (1999) (56). They were classified into three types (40 words for each): consistent (C: associate with only one pronunciation), inconsistent typical (IT: two or more possible pronunciations but the correct pronunciation is the most common), and inconsistent atypical (IA: the same as IT except that the correct pronunciation is in the minority) (Figure 5, A). As described for the IT and IA words, Japanese kanji words generally have one or more pronunciations. If a reader produces a pronunciation of a kanji character that is inappropriate for the specific target word but is legitimate for that character in other words of which it is a component, then such errors are referred to as legitimate alternative reading of components (LARC) (3, 57, 58). In general, the frequency of LARC errors reduces across word types in the order of: IA words > IT words > C words. As a control task, we also employed 56 non-word kana with 2 to 5 mora from the Sophia Analysis of Language in Aphasia (SALA) battery (Japanese version of PALPA: Psycholinguistic Assessments of Language Processing in Aphasia) (59).

**Figure 5.**
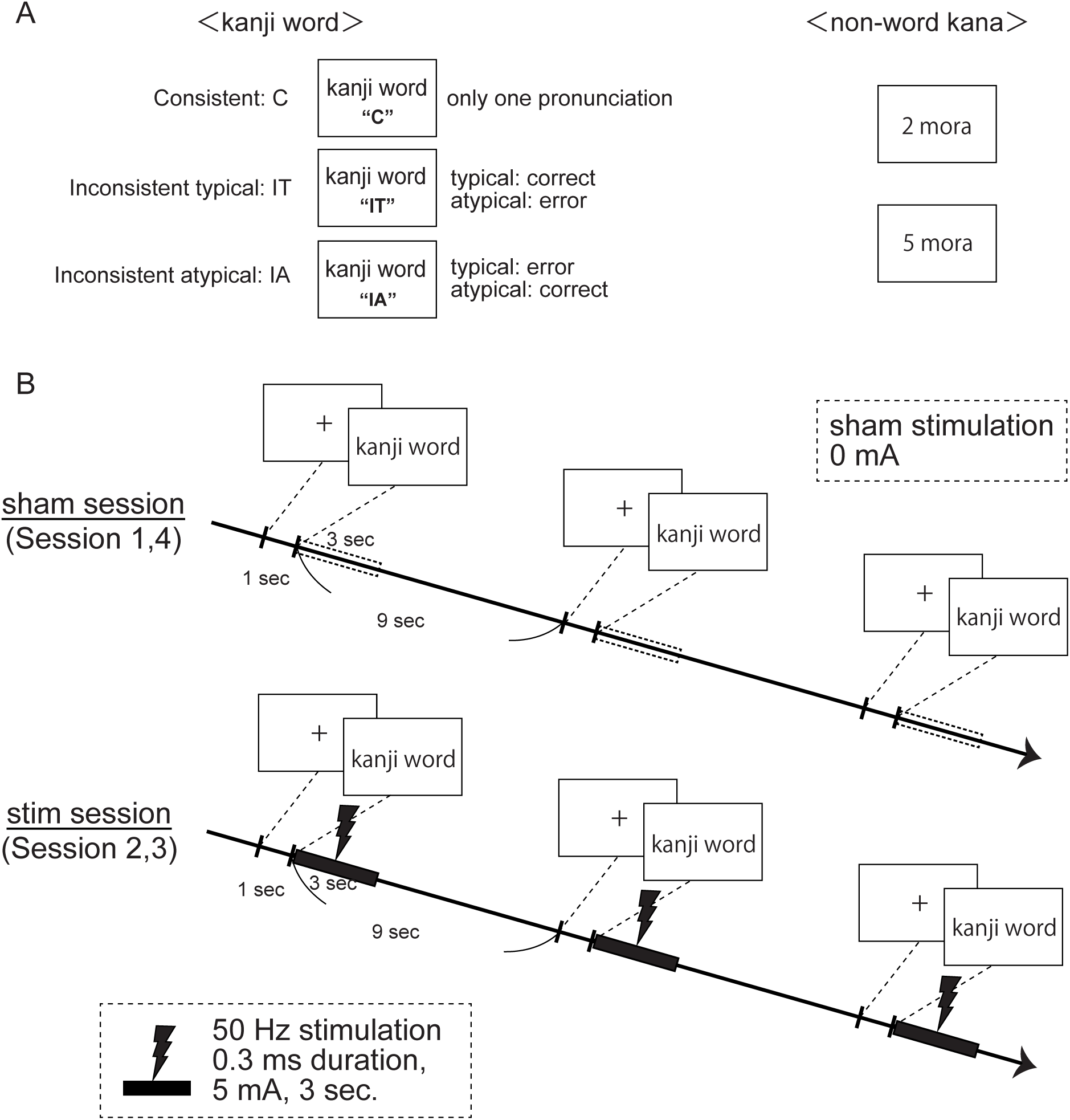
Stimulation protocol. A. Examples of items for reading task (Please contact the corresponding author for specific items). Each kanji word consists of two Chinese characters and is classified into three groups; Consistent word (C) which has only one pronunciation, Inconsistent typical word (IT) which has two or more possible pronunciations and the typical pronunciation is correct, and Inconsistent atypical word (IA) which is the same as IT word except that the atypical pronunciation is correct. We included 40 items for each group from Fushimi et al, 1999 (56). Non-word kana consists of Japanese kana with 2–5 mora, which make no sense (from SALA: Sophia Analysis of Language in Aphasia battery) (59). Each 2–5 mora non-word kana has 14 items. B. Stimulation protocol. The kanji reading task spanned four sessions followed by the non-word kana reading task over another four sessions. During session 1 and 4, sham stimulation (no stimulation) was delivered, and during session 2 and 3, real stimulation (50 Hz stimulation, 0.3 ms duration, 5 mA, 3 sec) was delivered. During the stimulation session for each task, the stimulation was delivered time-locked to the stimulus presentation. Each kanji or non-word kana was presented every 10 seconds including one second fixation.

Before subdural electrode implantation, all patients completed the kanji and SALA non-word kana reading tasks in order to confirm good baseline performance. Each item was presented individually on the computer screen, and we evaluated accuracy and reaction time (RT) by use of simultaneous photoelectric sensor and sound recordings for accurately detecting the timing of the presentation and answer, respectively. The patients were asked to read each word as quickly and accurately as possible.

In the tasks utilized to assess reading performance during subdural electrode stimulation, we used only the kanji and non-word kana items which each patient had correctly pronounced in the pre-operation study. We divided the items into four groups so that each group had the baseline RT, and assigned these four groups to two sham (without stimulation, “sham”) and two stimulation (“stim”) sessions. The four sessions (two sham and two stim sessions) were performed in order of sham-stim-stim-sham session to control for any changes in the patients’ attention (Figure 5, B). Each session had between 17–30 kanji items and 3–14 non-word kana items. The kanji reading task was completed over four sessions followed by the non-word kana reading task, across another four sessions. Patient 1 only completed the kanji reading task due to fatigue. As for the pre-operative assessment, all patients were asked to read aloud the kanji words and non-word kana, each presented individually on the computer screen every 10 seconds (including one second fixation). Again, we recorded the timing of the presentation of kanji words or non-word kana by photoelectric sensor and the timing of the answer by microphone, and then calculated the RT and evaluated the accuracy.

### Stimulation

#### Stimulus site

A magnetization-prepared rapid gradient-echo (MPRAGE) sequence was applied for anatomical T1-weighted volume data acquisition. An MPRAGE volumetric scan was performed before and after implantation of subdural electrodes as a part of presurgical evaluations. In the volumetric scan taken after implantation, the location of each electrode was identified on the 2D slices using its signal void due to the property of platinum alloy (60). To compare the findings obtained in individual patients with previous fMRI/PET semantic studies (the results of which are reported in standard space), the location of electrodes was coregistered to the presurgical 3D-MRI, and then normalized to the MNI standard space for anatomical localization. Electrodes identified on the T1 volume acquisition (1.5 T, MPRAGE) taken after grid implantation were non-linearly coregistered to the T1 volume acquisition taken before implantation (3T, MPRAGE), then to the MNI standard space (ICBM-152) using FNIRT of the FSL software (www.fmrib.ox.ac.uk/fsl/fnirt/). This method has been reported elsewhere for standardization of the electrode locations (61). The BTLA was defined as the cortical region, which exhibited marked, transient language impairment during electrocortical stimulation (in the clinically-performed functional brain mapping) (46). We defined the anterior part of basal temporal language area (BTLA) as falling within 6 cm of the tip of the temporal pole (in MNI space) and the remaining posterior part was defined as the posterior BTLA. This division was based on a previous cortical electrical stimulation study (30) which showed via electrical cortical stimulation that the “visual language area” is located in the middle third of the dominant fusiform gyrus, and this area is about 6 cm away from the temporal pole. In Patient 1, we stimulated only the anterior BTLA and she performed only kanji reading task in anterior BTLA stimulation due to fatigue. Patient 2, 3 and 4 performed both kanji and non-word kana reading tasks in anterior and posterior BTLA stimulation (Figure 2, A). In Patient 5, we stimulated the electrode pair located in the SMG instead of the posterior BTLA because no subdural electrodes covered the posterior BTLA. The SMG electrodes were selected according to the results of the functional cortical mapping (which showed a transient language impairment at this site), and the cortico-cortical evoked potentials (CCEP) which confirmed its connection to Broca’s area (see ref. for the CCEP method). Accordingly, we considered that this part of SMG fell within the dorsal language pathway and is a key region for phonological processing (35, 37, 62). Accordingly, Patient 5 performed both kanji reading tasks and non-word kana reading tasks in anterior BTLA and SMG stimulation (Figure 1, A).

#### Stimulation methods

High frequency electrical cortical stimulation (ECS) was performed through the subdural electrodes. ECoG was recorded at the sampling rate of 1000 Hz with a band-pass filter of 0.016-300 Hz (Patient 1–4) or 2,000 Hz with a band-pass filter of 0.016-600 Hz (Patient 5) to monitor for afterdischarges. Repetitive, square-wave electric currents of alternative polarity with a pulse width of 0.3 ms and a frequency of 50 Hz were delivered in a bipolar fashion through a pair of electrodes for 3 s (electrical stimulator SEN-7203, Nihon Kohden, Tokyo, Japan). The methodological details of the high frequency ECS for functional mapping have been described elsewhere (63). During the stimulation session for each task, the 50-Hz ECS was delivered time-locked to the presentation of kanji word or non-word kana with an intensity of 5 mA and duration of 3 s (Figure 5, B). In the functional cortical stimulation for clinical mapping, the intensity of stimulation is 10-15 mA in order to produce a binary (all or none) response outcome. However in this study, we used a lower (5 mA) intensity stimulation which can be used for quantitative RT analysis of the responses (46). Consequently, this stimulation did not induce afterdischarges. Exceptionally, in Patient 5, we used 3 mA for 3 s stimulation of anterior BTLA because stimulation at 5 mA caused facial twitch (current spread to the facial nerve) (64) and was also too strong for the naming tasks in clinical mapping procedure. For SMG stimulation of Patient 5, we used 9 mA for 3 sec stimulation because 5 mA stimulation was insufficient for quantitative RT analysis. In sham sessions, no stimulation was delivered. All patients could not recognize whether stimulation was given or not in each trial. After we finished all the tasks for anterior BTLA stimulation, we moved on to the tasks in posterior BTLA (Patient 1–4) or SMG (Patient 5) because it takes time to switch the electrode pair.

#### Semantic reliance (SR)

In the final part of this study, we also conducted a preliminary exploration of individual differences in reading, particularly with regard to SR (15). Following these recent studies, we operationalized the level of SR for each patient for the three types (IA, IT, C) of kanji word – by examining degree of correlation between item reading speed (from the presurgical baseline assessment) and imageability (a measure of the semantic richness for each word), partialling out the parallel effect of word frequency. Previous assessments of reading in English and Japanese (65, 66) have shown that the imageability effect increases from consistent-typically pronounced words (which can be read by the direct pathway) to inconsistent-atypical items (which require the additional input from semantics). People with high SR show enhanced effects of semantic variables such as imageability across all word types.

## Data Availability

All data produced in the present study are available upon reasonable request to the authors.

---

**Table SI-1.**
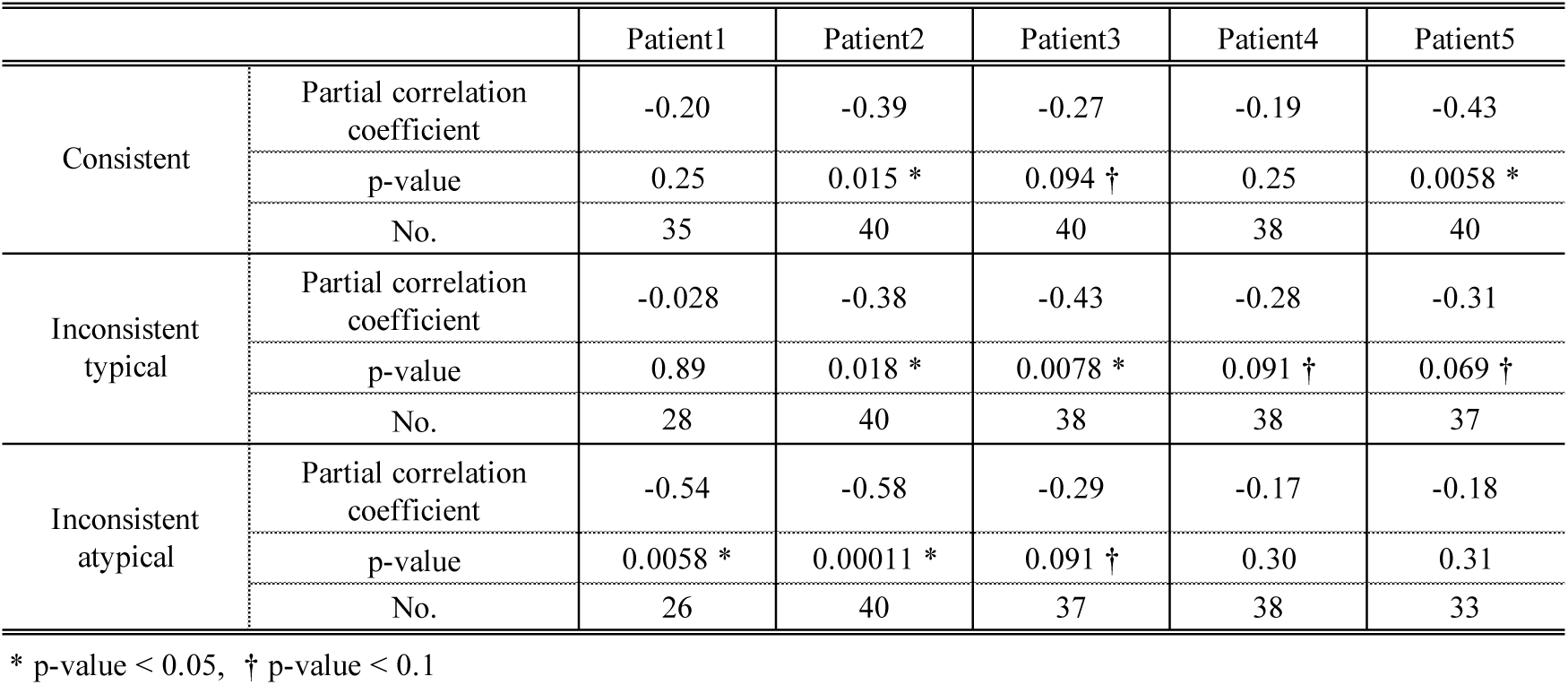
Semantic reliance for each patient.

**Table SI-2.**
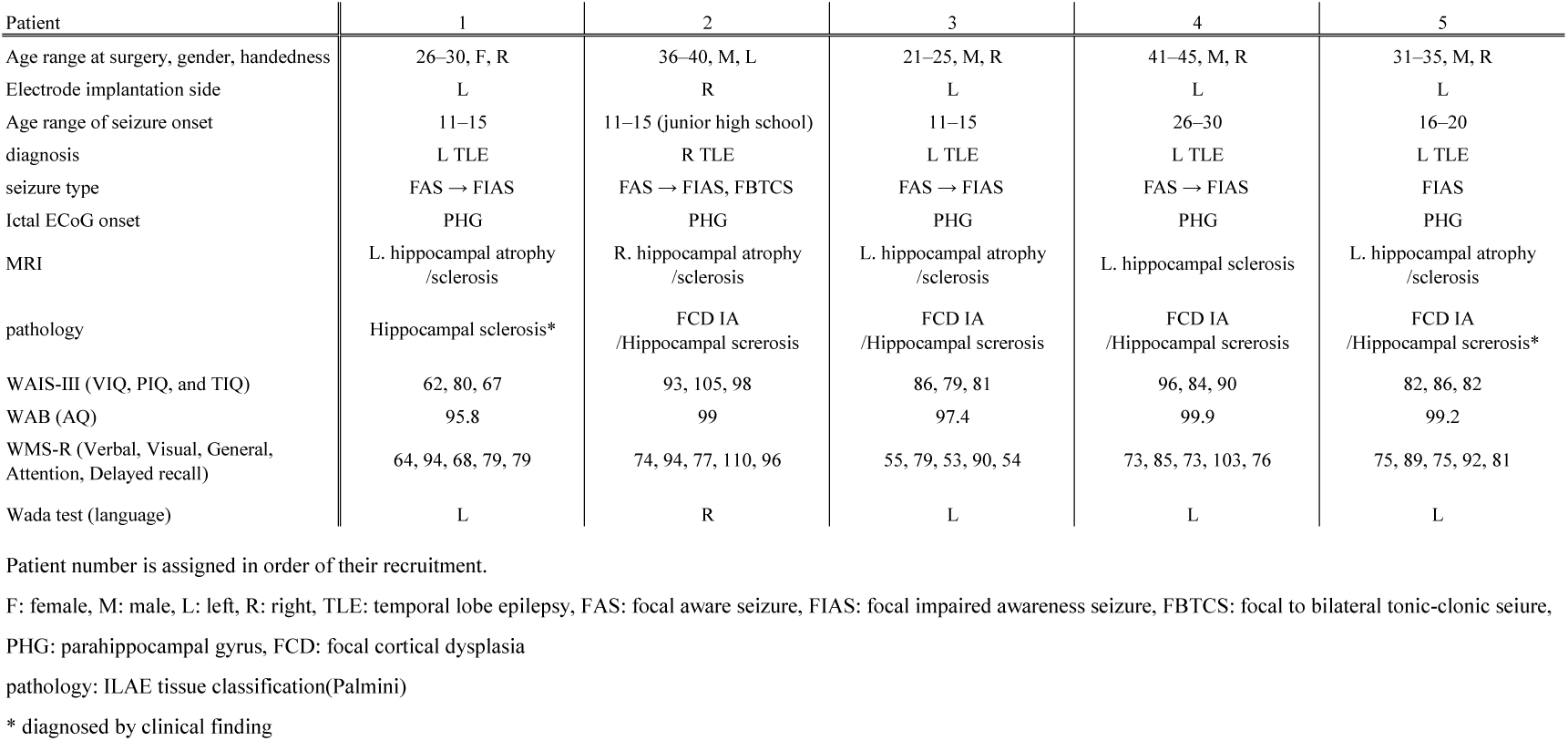
Patients’ demographics and clinical information.

