## Supplemental Table 1 for "Triangulating the neural cornerstones of reading: Within-participant double dissociations induced by direct cortical stimulation"

Table SI-1. Semantic reliance for each patient

| | | Patient1 | Patient2 | Patient3 | Patient4 | Patient5 |
|---|---|---|---|---|---|---|
| Consistent | Partial correlation coefficient | -0.20 | -0.39 | -0.27 | -0.19 | -0.43 |
| | p-value | 0.25 | 0.015 * | 0.094 † | 0.25 | 0.0058 * |
| | No. | 35 | 40 | 40 | 38 | 40 |
| Inconsistent typical | Partial correlation coefficient | -0.028 | -0.38 | -0.43 | -0.28 | -0.31 |
| | p-value | 0.89 | 0.018 * | 0.0078 * | 0.091 † | 0.069 † |
| | No. | 28 | 40 | 38 | 38 | 37 |
| Inconsistent atypical | Partial correlation coefficient | -0.54 | -0.58 | -0.29 | -0.17 | -0.18 |
| | p-value | 0.0058 * | 0.00011 * | 0.091 † | 0.30 | 0.31 |
| | No. | 26 | 40 | 37 | 38 | 33 |

* p-value < 0.05, † p-value < 0.1
