## Supplemental Table 2 for "Triangulating the neural cornerstones of reading: Within-participant double dissociations induced by direct cortical stimulation"

Table SI-2. Patients' demographics and clinical information

| Patient | 1 | 2 | 3 | 4 | 5 |
|---|---|---|---|---|---|
| Age range at surgery, gender, handedness | 26–30, F, R | 36–40, M, L | 21–25, M, R | 41–45, M, R | 31–35, M, R |
| Electrode implantation side | L | R | L | L | L |
| Age range of seizure onset | 11–15 | 11–15 (junior high school) | 11–15 | 26–30 | 16–20 |
| diagnosis | L TLE | R TLE | L TLE | L TLE | L TLE |
| seizure type | FAS → FIAS | FAS → FIAS, FBTCS | FAS → FIAS | FAS → FIAS | FIAS |
| Ictal ECoG onset | PHG | PHG | PHG | PHG | PHG |
| MRI | L. hippocampal atrophy /sclerosis | R. hippocampal atrophy /sclerosis | L. hippocampal atrophy /sclerosis | L. hippocampal sclerosis | L. hippocampal atrophy /sclerosis |
| pathology | Hippocampal sclerosis* | FCD IA /Hippocampal screrosis | FCD IA /Hippocampal screrosis | FCD IA /Hippocampal screrosis | FCD IA /Hippocampal screrosis* |
| WAIS-III (VIQ, PIQ, and TIQ) | 62, 80, 67 | 93, 105, 98 | 86, 79, 81 | 96, 84, 90 | 82, 86, 82 |
| WAB (AQ) | 95.8 | 99 | 97.4 | 99.9 | 99.2 |
| WMS-R (Verbal, Visual, General, Attention, Delayed recall) | 64, 94, 68, 79, 79 | 74, 94, 77, 110, 96 | 55, 79, 53, 90, 54 | 73, 85, 73, 103, 76 | 75, 89, 75, 92, 81 |
| Wada test (language) | L | R | L | L | L |

Patient number is assigned in order of their recruitment.

F: female, M: male, L: left, R: right, TLE: temporal lobe epilepsy, FAS: focal aware seizure, FIAS: focal impaired awareness seizure, FBTCS: focal to bilateral tonic-clonic seiure, PHG: parahippocampal gyrus, FCD: focal cortical dysplasia

pathology: ILAE tissue classification(Palmini)

* diagnosed by clinical finding
